# Exploring the role of the Care and Health Information Exchange (CHIE) in clinical decision-making: a realist evaluation

**DOI:** 10.1101/2020.08.13.20174276

**Authors:** Philip Scott, Elisavet Andrikopoulou, Haythem Nakkas, Paul Roderick

## Abstract

**Background:** The overall evidence for the impact of electronic information systems on cost, quality and safety of healthcare remains contested. Whilst it seems intuitively obvious that having more data about a patient will improve care, the mechanisms by which information availability is translated into better decision-making are not well understood. Furthermore, there is the risk of data overload creating a negative outcome. There are situations where a key information summary can be more useful than a rich record.

The Care and Health Information Exchange (CHIE) is a shared electronic health record for Hampshire and the Isle of Wight that combines key information from hospital, general practice, community care and social services. Its purpose is to provide clinical and care professionals with complete, accurate and up-to-date information when caring for patients. CHIE is used by GP out-of-hours services, acute hospital doctors, ambulance service, GPs and others in caring for patients.

**Research questions:** The fundamental question was “How does awareness of CHIE or usage of CHIE affect clinical decision-making?” The secondary questions were “What are the latent benefits of CHIE in frontline NHS operations?” and “What is the potential of CHIE to have an impact on major NHS cost pressures?”

The NHS funders decided to focus on acute medical inpatient admissions as the initial scope, given the high costs associated with hospital stays and the patient complexities (and therefore information requirements) often associated with unscheduled admissions.

**Methods:** Semi-structured interviews with healthcare professionals to explore their experience about the utility of CHIE in their clinical scenario, whether and how it has affected their decision-making practices and the barriers and facilitators for their use of CHIE. The Framework Method was used for qualitative analysis, supported by the software tool Atlas.ti.

**Results:** 21 healthcare professionals were interviewed. Three main functions were identified as useful: extensive medication prescribing history, information sharing between primary, secondary and social care and access to laboratory test results. We inferred two positive cognitive mechanisms: *knowledge confidence* and *collaboration assurance*, and three negative ones: *consent anxiety, search anxiety* and *data mistrust*.

**Conclusions:** CHIE gives clinicians the “bigger picture” to understand the patient’s health and social care history and circumstances so as to make confident and informed decisions. CHIE is very beneficial for medicines reconciliation on admission, especially for patients that are unable to speak or act for themselves or who cannot remember their precise medication or allergies. We found no clear evidence that CHIE has a significant impact on admission or discharge decisions.

We propose the use of “recommender systems” to help clinicians navigate such large volumes of patient data, which will only grow as additional data is collected.

## Introduction

One of the central goals of health informatics is improving care quality and safety by improving access to patient data^1, 2^. There have been widespread expectations among policy makers that this will produce significant cost savings^3^. Experience has shown that there is not a simple linear relationship between information availability and improved outcomes^4^, ^5^. The achievement of substantial financial benefits from electronic health records (EHRs) has also been elusive^6, 7^. The various moving and interacting parts that are involved form a complex socio-technical problem space^8-10^ that includes: the selection and interpretation of patient data in clinical decision-making; the design, interoperability and implementation of the numerous software tools required; the changes imposed upon practice and workload; and the broad range of healthcare processes and settings involved.

### Programme theory and rationale for evaluation

In April 2000, the Central Hampshire Electronic Records Development and Implementation Programme (ERDIP) was established and led to the creation of the Hampshire Health Record (HHR)^11^. In 2017, the HHR was renamed the Care and Health Information Exchange (CHIE) to reflect the facts that its scope had expanded beyond Hampshire to include the Isle of Wight and that it contained not only health but also social care data^12^.

The original purpose of HHR was to “test the usefulness of the electronic record in supporting emergency and out of hours care”^11^. CHIE now has a wide scope of application: usage data^13^ (Figure 1) shows there were 861,677 accesses in a 13-month period. The highest volume of use is from hospitals (44%), community and mental health services (28%) and general practitioners (GPs) (21%). There is also some use by ambulance services (2%), social services (1%) and “other” (2%).

**Figure 1:**
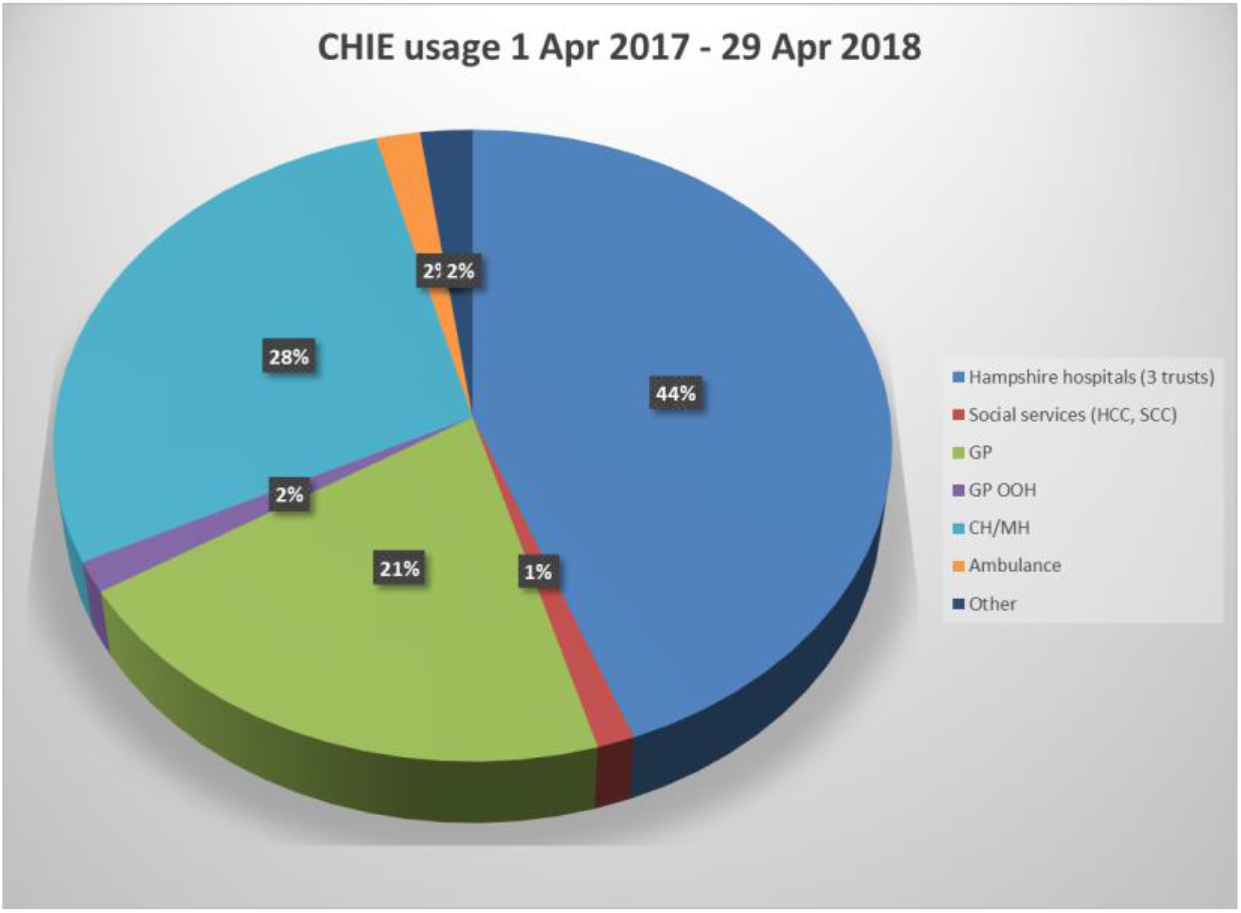
CHIE usage by organisation type.

The aim of CHIE is to provide “complete, accurate and up-to-date information for clinicians at the point of care”^12^, however the actual chain of causality from usage of CHIE to improved outcomes remained poorly understood. The “programme theory”, such as it is, is simply a general assumption that availability of linked patient data from primary care, secondary care and social care must be helpful. The purpose of this study was to open the “black box” of how clinicians used CHIE in practice and explore how it affected their decision-making.

At a time of great national interest in regional shared records (see Environment, in the Methods section), this study was conceived to offer learning from the longest-established of such UK projects to guide both national policy and implementation of new regional records. This study also aimed to lay the groundwork for a follow-up economic study to analyse the cost-effectiveness of CHIE: without some understanding of causal mechanisms, such an analysis would be impracticable.

### Evaluation questions, objectives and focus

The fundamental question was “How does awareness of CHIE or usage of CHIE affect clinical decision-making?” The secondary questions were “What are the latent benefits of CHIE in frontline NHS operations?” and “What is the potential of CHIE to have an impact on major NHS cost pressures?”

The funders decided to focus on acute medical inpatient admissions as the initial scope, given the high costs associated with hospital stays and the patient complexities (and therefore information requirements) often associated with unscheduled admissions. Specifically, the four use cases of interest were:

1. The decision to admit (Emergency Departments and Acute Medical Units).
2. Medicines reconciliation^14^ on admission (Acute Medical Units, general medical and elderly medicine wards).
3. Test ordering patterns for adult acute medical inpatients (Acute Medical Units, general medical and elderly medicine wards).
4. Discharge processes (Acute Medical Unit, general medical and elderly medicine wards).

## Methods

### Rationale for using realist evaluation

The UK Medical Research Council guidance^15^ defines process evaluation of complex interventions as “a study which aims to understand the functioning of an intervention, by examining implementation, mechanisms of impact, and contextual factors”^16^ and cites realist evaluation as an important theory-based approach for this kind of study. There is no doubt that implementation of a shared health and care record is a “complex” intervention: it comprises multiple interacting components ^15^ and faces numerous barriers to adoption^17^. It is exactly the “process” that we wished to understand: *how* does CHIE make a difference?

The structure of this paper is based upon on the RAMESES II guideline for realist evaluations^18^.

### Environment

Current NHS England health policy is described in the Long Term Plan^19^ and its information strategy centres around “global digital exemplars”^20^, interoperability standards^20^ and regional information sharing through “local health and care records”^21^. England is divided into 44 “sustainability and transformation partnerships” (STPs)^22^ to deliver the policy aims of integrated care. Hampshire and the Isle of Wight together constitute one STP, with a population of 2 million and four acute hospital groups^23^: University Hospitals Southampton NHS Foundation Trust (UHSFT), Hampshire Hospitals NHS Foundation Trust (HHFT), Portsmouth Hospitals NHS Trust (PHT) and Isle of Wight NHS Trust (IWT). The digital programme in the STP plan depends upon an integrated digital health and care record^24^.

Table 1 shows the staff and inpatient characteristics of the four HIOW hospital groups in the STP, based upon NHS activity reports for 2017-18^25, 26^ and workforce statistics for April 2018^28^, when this study was initiated.

**Table 1:** Participating hospital characteristics (FTE=full-time equivalent; HCP=qualified healthcare professional)

| Organisation | FTE HCP Staff | Admissions | Admissions to FTE staff ratio | A&E attendances | A&E attendances to FTE staff ratio |
| --- | --- | --- | --- | --- | --- |
| UHSFT | 5,471 | 148,840 | 27.2 | 141,530 | 25.9 |
| HHFT | 2,757 | 105,160 | 38.1 | 124,945 | 45.3 |
| PHT | 3,678 | 137,780 | 37.5 | 148,885 | 40.5 |
| IWHT | 1,335 | 28,105 | 21.0 | 50,435 | 37.8 |
| **Total** | **13,242** | **419,885** | **31.7** | **465,795** | **35.2** |

### Product

CHIE is fundamentally an information repository rather than an active decision-support system for a particular condition. It aggregates^27^ selected patient data from GP systems (only coded data, not free text) and (with some exceptions) correspondence (such as discharge summaries and outpatient clinic letters) and diagnostic reports (principally laboratory results and radiology reports) from the acute hospitals listed above and some social care data from Hampshire County Council and Southampton City Council. CHIE also includes correspondence from Care UK, who operate various services contracted out by the NHS within HIOW, and Royal Bournemouth and Christchurch Hospitals NHS Foundation Trust which makes referrals into HIOW services. CHIE has mental health data from Solent NHS Trust and Southern Health NHS Foundation Trust where the patient has given explicit consent.

CHIE does not rely on patient consent as its legal basis to process data under the General Data Protection Regulations (GDPR), but rather relies upon the legal provisions for ‘necessary protection of vital interests’ and ‘compliance with a legal obligation’^28^. It is still best practice to obtain patient consent wherever possible. Citizens can opt out of CHIE.

When the study protocol was designed, hospital usage of CHIE was split between HHFT (n=225,830 accesses between Apr 2017-Apr 2018; 59% of total hospital usage), UHSFT (n=106,883; 28%) and PHT (n=48,515; 13%). This wide variance reflected several underlying factors. Table 1 suggests significant differences in organisational workload, albeit from a very crude comparison. Each hospital had different ways of integrating access to CHIE with their local information systems, some with “single sign-on” to facilitate moving from a local application to CHIE. There had been varying levels of support from the clinical leadership in each of the hospitals. PHT in particular had a legacy of low usage due to a long period when the dominant GP systems supplier in the area had refused to submit data to CHIE, thus leading to a very negative perception by some clinicians. By the time of this study, CHIE data content from GP and hospital systems was very comprehensive. The major gap was diagnostic reports from HHFT. Nonetheless, pessimism about its value remained in some quarters. When this project began, CHIE had only recently become available in IWT so they were not included in the study design.

### Evaluation design

The evaluation programme was informed by our previously reported scoping review^29^. As our aim was to uncover the mechanisms by which CHIE facilitated improved outcomes, we decided to concentrate on recruiting clinicians who were frequent users.

We recruited a patient and public involvement (PPI) group to advise the project. Nine people attended a PPI meeting on 14 February 2018 to discuss the project and give formative advice. On 9 March 2018, the study design was discussed with the Young Adult Patient and Public Involvement (YAPPI) group organised by the South Central Research Design Service of the National Institute for Health Research (NIHR).

Based upon the research questions, we designed a semi-structured interview outline, an information sheet and a consent form and sought email feedback from our PPI group and the project steering group, which included a general practitioner, a hospital clinician and leaders of the STP digital programme. The final interview outline is given in the appendix.

### Data collection

We conducted single semi-structured audio-recorded interviews with healthcare professionals who were frequent users of CHIE, using a mixture of telephone and face-to-face interviews to obtain rich qualitative data while respecting the operational challenges of the highly pressurised healthcare services that our target participants worked within.

### Recruitment and sampling

We aimed to include junior doctors, consultants and pharmacists who worked in emergency departments, acute medical admission units, general medicine wards and elderly medicine wards. We identified frequent CHIE users in each hospital from the system usage logs, starting with the top 50 and working down the list. We emailed a flyer to them, explaining the project and inviting them to complete an online consent form. Once the consent form had been completed, we contacted the person to schedule a suitable time and place for the interview. Participation was re-confirmed at the start of each interview. We aimed to recruit up to forty participants. It is not possible to predict a number for qualitative saturation, but this seemed a reasonable goal based upon our scope and methods, taking account of prior experience^30^. Due to very low response to the emails, we later resorted to direct approaches to known enthusiasts by the digital programme leaders in each hospital.

### Data analysis

Interviews were audio recorded and later transcribed by a commercial service provider. The Framework method^31^ was used for qualitative analysis, which involves a systematic process of sifting, charting, interpreting and sorting material according to the project’s key issues and themes. This method is recommended for interview analysis^32^. We used Atlas.ti version 7 to support and document our qualitative analysis^33^.

Codes were allocated to recurring concepts in the transcripts and interconnections within the transcribed texts were identified. The themes were iteratively revised multiple times to accommodate all the codes and create the related subthemes. The grouped codes and transcribed text were analysed by identifying the characteristics and differences between the data, generating typologies, interrogating theoretical concepts between categories and sub-themes to explore relationships and causality.

### Ethical review

Our study protocol was submitted via the Integrated Research Application System (IRAS) on 4 April 2018 (ref 244805) and booked for Health Research Authority (HRA) review and proportionate review by an NHS research ethics committee. HRA subsequently decided that ethics committee review was unnecessary and granted approval on 16 May. The protocol was then submitted to the research governance process of the three participating hospitals. Approval to proceed was received from UHSFT on 23 May. Various questions arose from the other two hospitals during June and July, resulting in the need to submit an HRA amendment to extend the study end date from 31 July to 31 October. HRA approved the study extension on 31 July. HHFT gave approval to proceed on 6 August. PHT raised further queries but finally gave approval to proceed on 25 September.

## Results

### Participants

We interviewed 21 healthcare professionals from PHT (n=4), UHS (n=7) and HHFT (n=10). The participants consisted of physicians (n=14), nurses (n=6) and one research fellow. Given the small sample size we have not attempted to report further differentiation regarding the profession and specialty of the participants.

### Main findings

We identified three principal functions of CHIE and five cognitive mechanisms. The common context was inpatient healthcare provision by experienced and frequent users of CHIE.

### Function 1: Extensive medication prescribing history

Overall the extensive prescribing history from primary care assists healthcare professionals in minimising medication errors and make more informed decisions. A holistic picture of the extensive medication prescribing history mechanism, with both benefits and mitigation instances is shown in Figure 2. The following quotations illustrate this theme.

> P06: “….*I look to see what the GP’s prescribing them, so what their regular medications are and also to look back at the medication issues to see what they have been prescribed recently by their GP…”*

**Figure 2:**
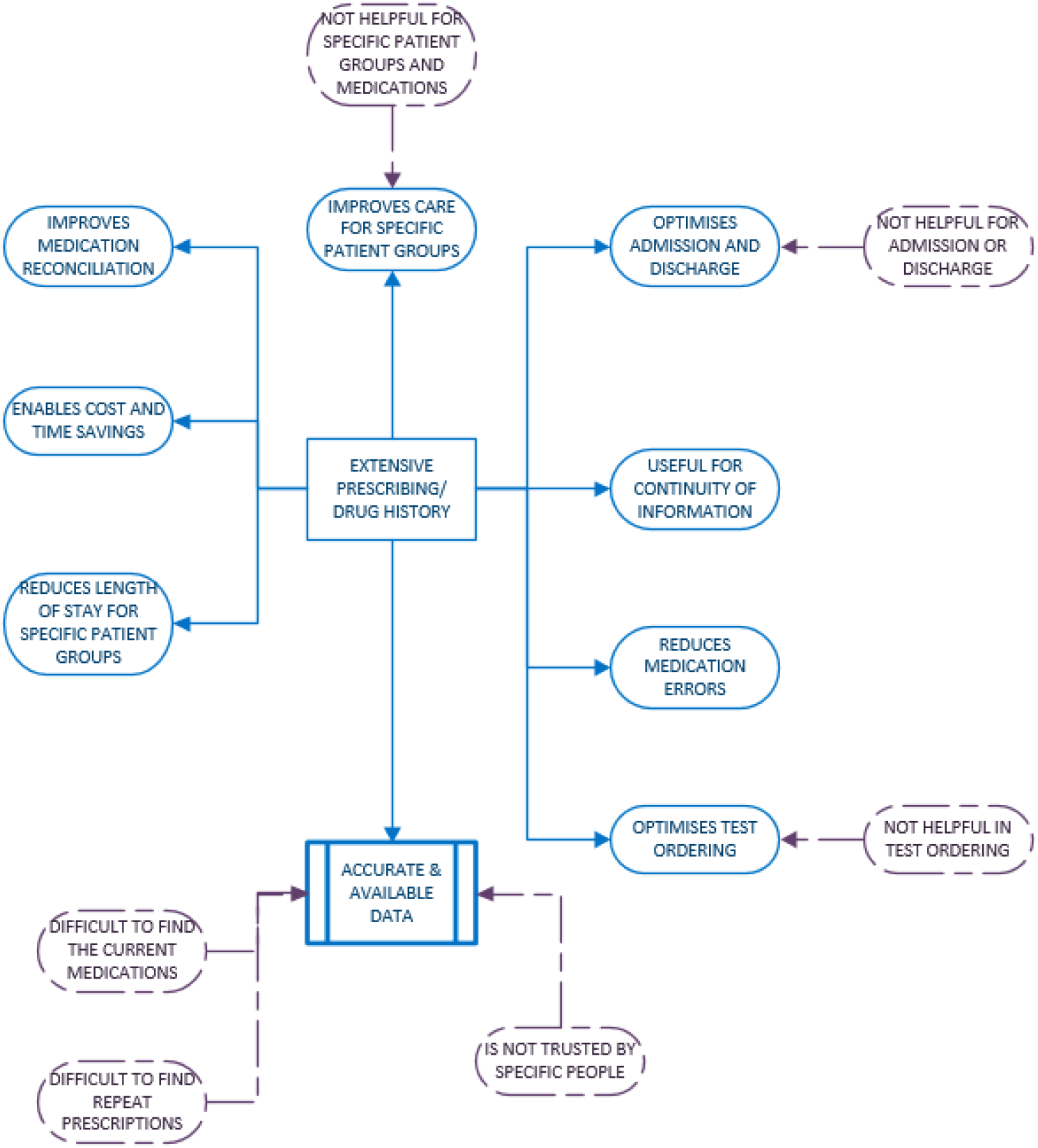
Function 1 - Extensive medication prescribing history.

The extensive medication prescribing history is especially helpful for people with long-term conditions or older and confused patients, as shown in these example comments.

> P02: “…*Migraine and epilepsy… with a comprehensive healthcare record…you can certainly look back ten years and see the medications that have been prescribed and that's extraordinarily useful when patients understandably can't remember what medication they tried previously*…”
>
> P13: “…*the CHIE enables me to note down what medication they're on and be able to prescribe it for them in advance if applicable for the next day if they're being admitted or ensure that they are discharged on the correct medication*…”
>
> P30: “…*I checked his regular medicines on CHIE against what we were prescribing him in hospital to make sure that he was still receiving the same drugs*…”

CHIE is also useful for the admissions and discharge processes and it may be able to reduce the length of stay. This might be possible because of the record of continuous information regarding the medication an inpatient is receiving in primary care or a patient that is about to be admitted. This record includes both hospital and GP records and this is what makes it uniquely useful to healthcare professionals and enable them to make informed decisions minimising their mistakes and improving patient care.

> P04: “…*it's like a checking mechanism for medication*…”
>
> P06: “…*for inpatients, we use the CHIE when they first come in and it improves the continuity of information from the GP record to the hospital record and so it reduces time caused by any confusion…it will reduce a hospital stay a little*…”
>
> P13: “…*it could potentially reduce the length of stay in you being able to see what the GP can provide in primary care and liaise with them in that way so you could have a look on healthcare records*…”
>
> P13: “…*clerking patients, especially getting a drug history from them because you can get an up to date GP certified list of their medications*…and allergies…”

The CHIE data are mostly accurate and available. This again has the same benefits of improving patient care, by minimising potential medication errors and saving time in enquiring about a patient’s history.

> P14: “…*It's made my life so much easier because all the information is up to date, so we don’t go duplication prescribing or trying new drugs that have already been tried*…”
>
> P15: “…*because sometimes some information we have is incomplete, so I use CHIE just to get the data about the patient which is not available for me from the patient who is sitting in front of me or from the computer systems here in the hospital*…”
>
> P02: “…*the most accurate information is what was prescribed by the GP, that's why I find it so useful*…”

The extensive medication history also provides a historical record of what has happened to a patient before. This is valuable knowledge especially when a patient is unable to communicate or in the common scenario that the patient’s relatives will not always have the best idea or remember everything on what has happened to their loved ones.

> P28: “…*you get a lot more evidence than if you’d tried to speak to the patient*…”

Perceived problems with patient consent cause the system to not be trusted by everyone and also to have missing and outdated patient data, since people opt in or out and they do not remember whether they did or not necessarily understand why CHIE is important for them. The lack of education of the patients and the healthcare professionals is the main point of reference for this problem.

> P29: “…*certain people don’t trust it, I would imagine… we got this email through saying, ‘You need to sign up for this to get access to it’ but actually I didn’t know what that was until a few months down the ine*…”

### Function 2: Information sharing between primary and secondary care

A holistic picture of the information sharing function, with both benefits and mitigation instances is shown in Figure 3.

> P06: “…*Definitely quality improvement…because it allows the continuity of information particularly with investigations, correspondence and medications and so that it helps reduce medication errors*.…”

**Figure 3:**
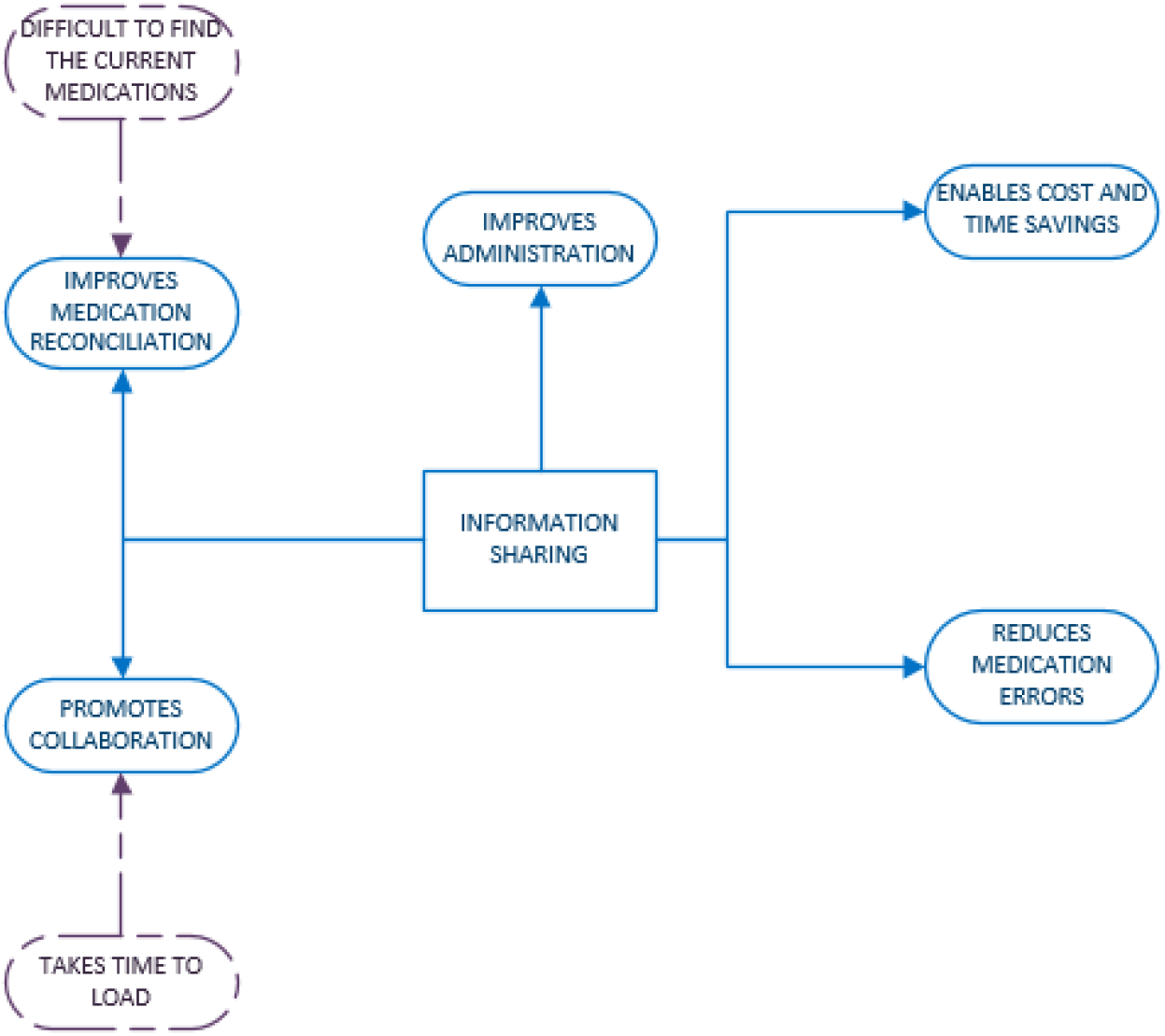
Function 2 - Information sharing between primary, secondary and social care.

Participants feel that information sharing is able to reduce workload and enable time and cost savings. It also enables healthcare professionals to not only make more informed decisions but also to feel more confident about their decisions and personalise care.

> P13: *“*…*it shares information without much of a work burden between primary, secondary and tertiary centres*…”
>
> P18: *“*…*see the background of the patient, reassure myself about the circumstances, and then phone the patient and email the GP to manage the situation so the patient doesn’t need to come in at all*.…*”*

Time is saved having this type of centralised system and it also acts as validation for what a patient remembers and what can be confirmed from the patient record. This shared information is also pre-emptively informing the healthcare professionals about relevant changes in medication or procedures that again a patient may find hard to remember.

> P14: “…*acts as like a useful tool to check the information the patient has, what’s actually happening, what’s been requested*.…*”*
>
> P16: *“*…*you can get more information than you can otherwise from a patient, or expecting a patient to call you to tell you about a change. You can already see what happened*.…*”*

This validation is able to sometimes reduce over-prescription, prevent medication errors and improve medication reconciliation, since the prescriber can be sure of the regular medication a patient takes.

> P17: “…*you can clarify what their normal prescriptions are so you’re not over-prescribing*.…*”*
>
> P31: “…*safeguarding point of view because we can see what has been prescribed recently*.…”
>
> P35: “…*optimise patients’ medication, having access to their out of area blood results…”*

The shared information is especially useful for outpatient clinics and assists with the decision making whether a patient will be admitted.

> P18: *“*…*when I’m asked for advice on patients, whether that be patients in the hospital or patients outside the hospital*…”
>
> P32: *“*…*It just helps to see it in one snapshot*…”

This shared information enables the healthcare professionals to have a greater picture of the patient history, containing measurements about blood pressure and height, which are typically done in primary care.

> P32: “…*I found it very useful to look at baseline physiology…it’s most useful for blood pressure recordings, because GPs tend to do that, which would be that they recorded oxygen saturations more reliably*…”

CHIE is also quite helpful from a social background and administration point of view because it contains information such as letters to GP, clinic consultation, admissions and administrative details. Administration errors are easily identified and minimised.

> P15: “…*sometimes patients will say I’ve been referred but I cannot see anything on our system because the referral has not been received yet*…”

CHIE promotes efficient and effective work collaboration between hospitals to improve patient care, outcomes and treatment and clinical decision making and therefore might be able to reduce the length of stay.

> P31: “…*being able to look up investigation results that have been done in Southampton and I found that most useful for the patients I manage jointly alongside the specialists in Southampton…”*
>
> P32: “…*a lot of patients sit between a number of different trusts, and CHIE is very good at pulling together…discharge summaries, different requirements*…”
>
> P04: “…*It doesn’t necessarily help me whether to decide to admit a patient but.it would make a difference potentially to the treatment decisions that I make*…”
>
> P33: “…it may reduce the length of stay…*more information available to make clinical decisions*…”

A relevant problem is that although the data are not necessarily missing or outdated, the format or the details included in them are such that make them nearly unusable. There have been very few instances of missing data and this is predominantly because of the locality of the CHIE. The missing, outdated or badly formatted data cause problems and minimise the positive outcomes of the system. This causes the system to not be as helpful for specific group of patients, not being helpful in test ordering and not seen as helpful for admission and discharge processes.

> P02: “.no *structure to the results being uploaded and the reference intervals aren’t included… it doesn’t allow us to look into the GP records in granular detail*…”
>
> P03: “. *it doesn’t go into enough granular detail about events*…*”*
>
> P06: “. *the problem is, its investigations are often incomplete*…*”*
>
> P33: “…*not up-to-date and it hasn’t got all the information on it, that can be frustrating…”*

There were many comments that CHIE needs to be more user-friendly and more technically efficient.

> P33: *“*…*I guess if it’s not up-to-date and it hasn’t got all the information on it, which can be frustrating*… *The layout used to be much more user-friendly, whereas the new format is not so user-friendly…”*
>
> P03: *“*…*I can’t get any information, either there’s some sort of IT error or the information isn’t there for some reason…”*
>
> P02: “…*the CHIE records are clearly out of date so the patient is on medication but the last recorded prescription is six months earlier*…”

CHIE has further technical difficulties such as complicated navigation functionality and it is considered hard to find where all the information is. These problems could arguably be due to lack of training or low IT literacy of the healthcare professionals. However, it could also possible that these problems are due to the lack of awareness of the type of functionality exists and also due to very limited time the healthcare professionals have when visiting a patient.

> P16: *“*…*I’m yet to find where the pharmacy information is*…”
>
> P14: *“*…*our IT system doesn’t always function 100% of the time*…*”*
>
> P02: *“*…*we haven’t in any way empowered the workforce to use it properly. So that’s a failure of IT and of the training programmes that we put onto people when they start a new role*…*”*

### Function 3: Viewing laboratory results

The ability to view a variety of laboratory test results further enhances the complete picture a healthcare professional has about their patient. A holistic picture of the viewing lab results mechanism, with both benefits and mitigation instances is shown in Figure 4.

**Figure 4:**
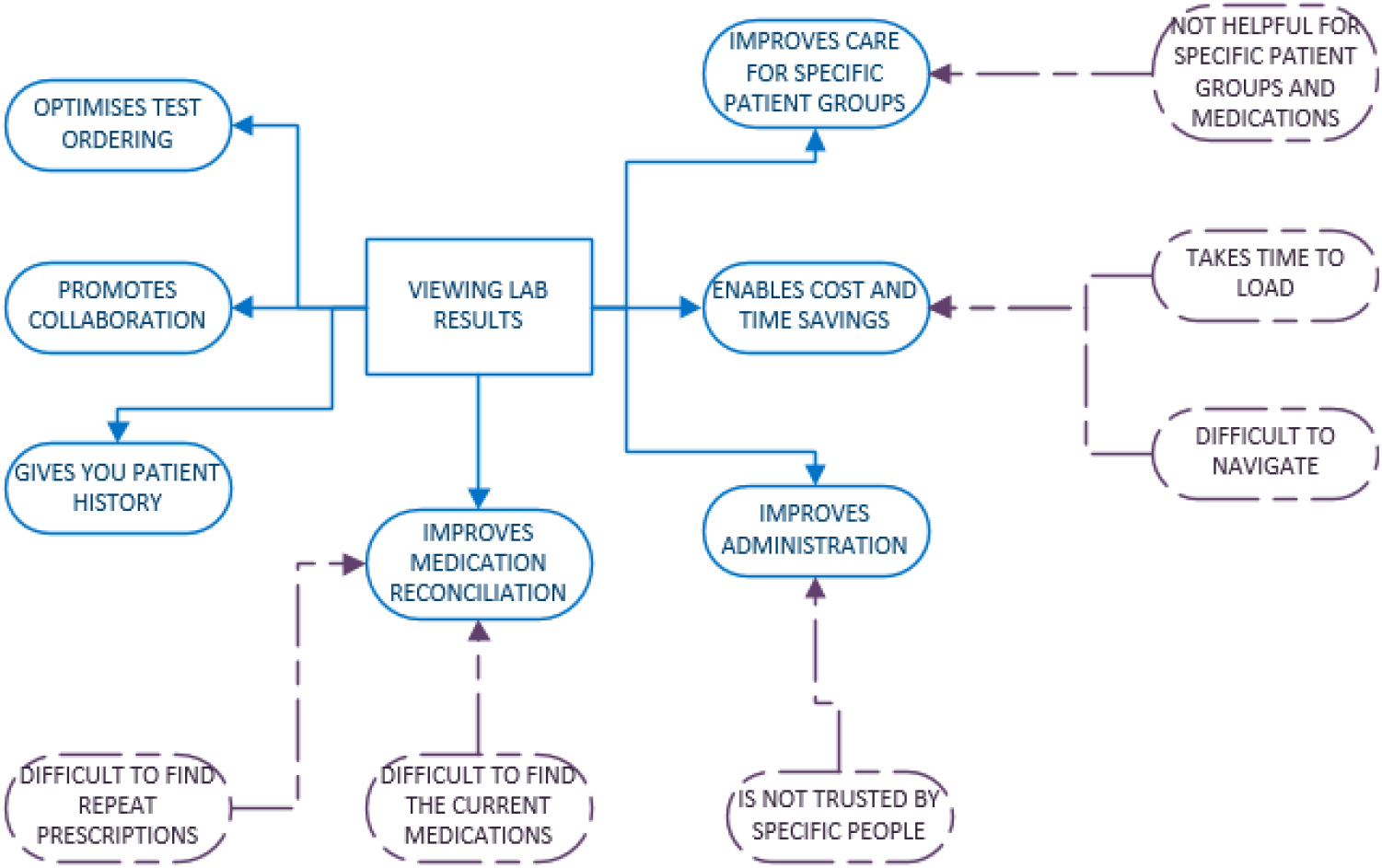
Function 3 - Viewing laboratory results.

This information is important to ensure that the patient’s care is managed in the most timely and effective manner.

> P01: “…*looking at results of blood tests and looking at what medication they’re currently prescribed… affects how I manage their care*…”
>
> P05: “…*I just go to the CHIE, look up their results through the system rather than having to ring the lab…*”

It is felt to promote efficient and effective work collaboration between trusts to improve patient care.

> P02: “*…able to see blood test results from other labs outside of UHS so Portsmouth bloods or Hampshire hospital bloods, that’s very useful*…”

A common problem a lot of healthcare professionals face is the compartmentalised records of patients. Many patients are seen in several NHS organisations. CHIE includes a laboratory results and medication view that assists diagnosticians and frees administration staff from making phone calls to several laboratories or GPs to actually perform their other duties in a more efficient manner.

> P05: “*…really useful diagnostically because I may only have a small pocket of results in our system if somebody happens to live in Southampton, but I get referred patients from Portsmouth and Basingstoke and Winchester and all around and having no results makes is really hard. So what I used to have to do was to write to GPs or get my secretary to ring up and then because they may not have got the same medical background as me, the results that would come back may not be quite as informative. So, having those results, it’s really drug record and results are the two big things that improve patient care*…”
>
> P34: *“…patients coming from out of area, if their information is on CHIE, that’s very useful*…*”*

The laboratory tests are also act as a validation mechanism to verify the historical records of a patient or enhance their perception of their health based on what they remember.

> P14: *“…when bloods have been done, we can often find out information, whether the patient has had the information or not*…*”*

The accessibility of current and old laboratory test results, prevent duplication of tests, saving further time and improves medication reconciliation and assists in decision-making for further tests.

> P02: “*…reducing duplicate investigations*…*”*
>
> P05: “*…there’s some tests which they can’t process on the same day, but actually it has already been done, and you without a doubt, you’re going to save time, so it’s about preventing duplication*…*”*
>
> P06: “*…I think it really improves patient safety in terms of medicines reconciliation and it reduces duplication of care and investigations*…*”*
>
> P33: “*…like an echo report, it means that you wouldn’t have to repeat that test*…”

However, the system has caused concern when some of the data were outdated. These outdated data were often caused because the patients were seen in a different hospital that is not using CHIE. The cause for the data being outdated or seemingly missing could vary greatly from a human error, to the system being too slowly updated from source systems.

> P02: *“…pharmacy to you is one example.the CHIE records are clearly out of date so the patient is on medication but the last recorded prescription is six months earlier*…”
>
> P04: “*…They’re also not always up to date so if I go in to look for a medication history, it might be several years old*…*”*
>
> P01: “*…Sometimes the information doesn’t get there quickly enough for me to see it*…*”*

### Aspirations

Participants would like CHIE to assist them deciding what kind of procedures to do and whether to discharge or admit a patient.

> P15: “*…*we *don’t have a system where we make a decision about CPR*…*”*

Some would like for all the organisations in the area to have a common shared system and they acknowledge the need for awareness on CHIE benefits and usage boost access.

> P01: “*…if all of the care is based within a hospital setting they possibly don’t think to look elsewhere*…*”*
>
> P30: “*…I’d hope that in the future more thought and work will be put into that side of things, as well as getting more providers on board with uploading information and sharing it with other people*…*”*
>
> P03: *“…ultimately a joined-up IT system across the region is absolutely the correct direction of travel*…*”*

Registration requirements and the speed of the system may need to be more efficient and less complicated. Also added functionality could improve the effectiveness of the use and also the time a healthcare professional spends using it effectively; in comparison to the time spent using the CHIE system desperately trying to find information that although it is there, it is not easily accessible.

> P05: “*…not having the searchable capability for results*…*”*
>
> P15: “*…it would be much easier if there was just like a list of current medications*…”
>
> P32: “*…I think it would be even better if it was an electronic system for accepting GP referrals real-time, and the ambulance EPR in real-time*…*”*

Further training is needed for the healthcare professionals and the patients, on how to use the CHIE system and why it is beneficial for the patients to consent for the system to be used for their data.

> P02: “*…patient education to make sure the patient understands what the purpose of this system is and why actually the data needs to be granular*…”
>
> P14: “*…I think it’s about education….we’ve always sent a copy of our letter to the patient and that has been so important because then there’s no agenda, there’s no underlying concerns by the patient that we’re keeping things from them*…*sometimes they don’t like what they read in the letters because that isn’t them but actually it’s really helpful for patients to see how they’re perceived by other people and help them with their management as well*…*”*
>
> P13: *“…a bit of training on it would be useful. I don’t want to increase too much workload, but just to understand the different aspects of it now, especially on this new interface…”*

## Discussion

Our study has identified three functions by which CHIE can achieve the desired outcome of improved care in the context of three NHS hospitals: the extensive medication history; the information sharing between primary, secondary and social care; and viewing previous laboratory tests. These functions are specific features of the CHIE “intervention”. “Mechanisms” are defined in realist evaluation as “underlying entities, processes or structures which. generate outcomes of interest”^11^ or “a combination of resources offered by the social programme under study and stakeholders’ reasoning in response”^12^.

Grouping together how the three functions are used, we infer two inter-related cognitive mechanisms: *knowledge confidence* and *collaboration assurance*. Knowledge confidence is the clinician’s sense that they have sufficient (and sufficiently reliable) information for the decision at hand or that their initial thinking has been suitably modified. Collaboration assurance is the warranted belief that other healthcare providers are in fact fulfilling their part of shared patient management.

We also infer three mechanisms which work against realising the intended benefits. Apart from poor general awareness of the existence and contents of CHIE (determined anecdotally), there is apparently further need for clear and plain language explanation about the legal basis for accessing CHIE. We label this *consent anxiety*. Other barriers were the perceived difficulty of finding the desired nuggets of data in a sea of irrelevance; we call this *search anxiety*. The third element is a cognitive bias introduced by bad experiences with finding incorrect or missing data in CHIE – this is *data mistrust*.

Figure 5 depicts the interaction of CHIE as the intervention, with its three key functions, and the various cognitive mechanisms which enable or hinder the desired outcomes.

**Figure 5:**
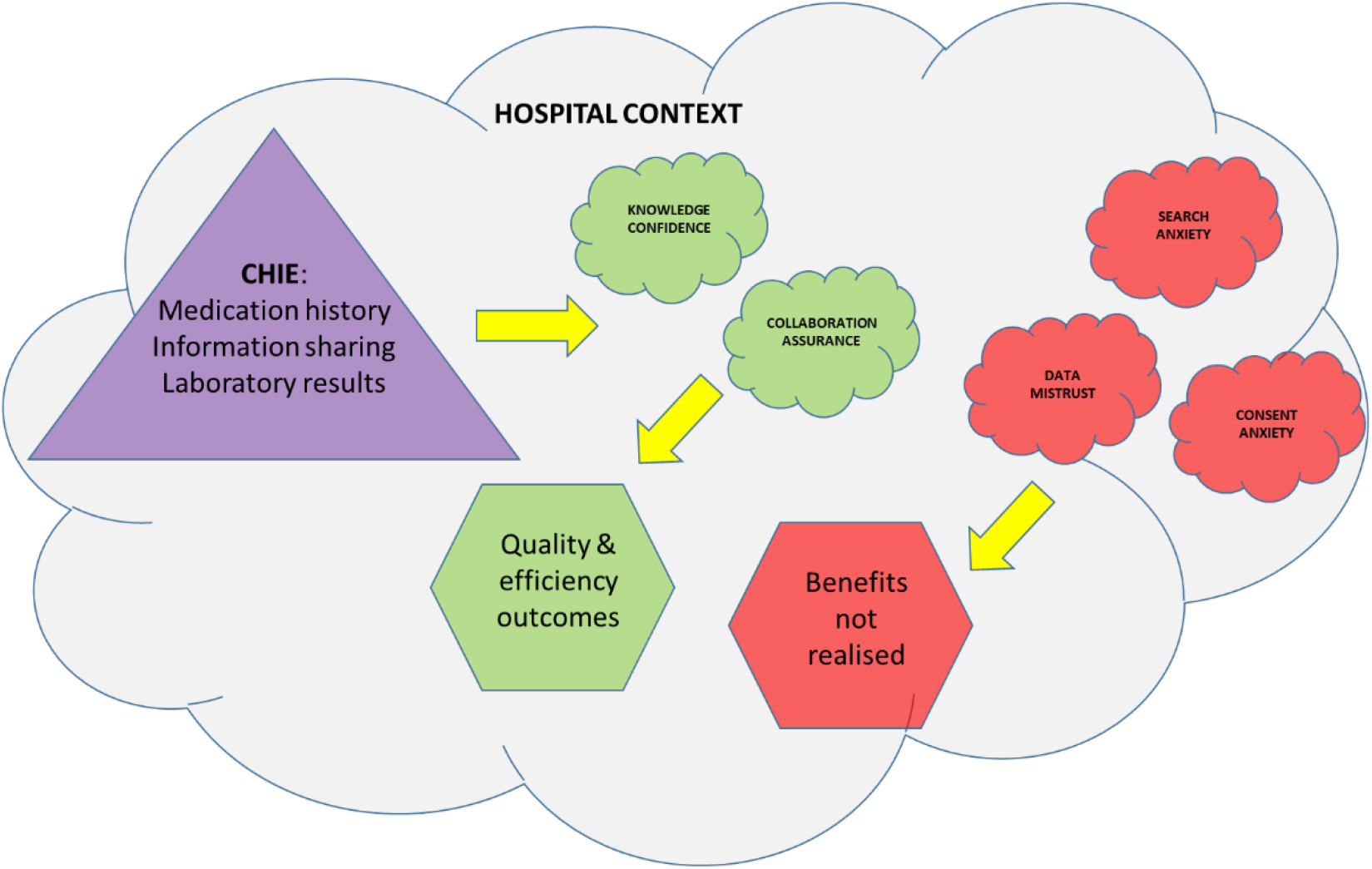
Context and mechanisms for realised outcomes.

### Evaluation of primary research question by use case

| Use case | How does awareness of CHIE or usage of CHIE affect clinical decision-making? |
| --- | --- |
| Decision to admit | We found no particular impact on admission decisions in general, but CHIE does give clinicians the “bigger picture” to understand the patient’s health and social care history and circumstances so as to make confident and informed decisions. In some cases, knowledge of previously investigated diagnoses helped to decide whether to admit. |
| Medicines reconciliation on admission | We found that CHIE is very beneficial for medicines reconciliation on admission, especially for patients that are unable to speak or act for themselves or who cannot remember their precise medication or allergies. The effect was in the timeliness and efficiency of decision-making. |
| Test ordering patterns | CHIE definitely helps clinicians decide when an intended test can be avoided if previous results are found. However, it is not considered to be helpful for deciding which test should be requested in the case at hand. In some cases, the information on CHIE is incomplete, outdated or insufficiently granular so some tests may still be duplicated. |
| Discharge processes | We found no clear evidence that CHIE has a significant impact on discharge decisions. |

### Secondary research questions

What are the latent benefits of CHIE in frontline NHS operations? For acute medical inpatients, the primary impact seems to be in medicines reconciliation and test ordering. There does not seem to be a strong case to anticipate significant impact on admission or discharge decisions as so many other factors are involved.

The latency of the benefit is largely down to confounding by indication, as noted in our scoping review^29^, and lack of awareness of what CHIE offers, how it is accessed or what the information governance rules are about its usage. Some technical barriers were highlighted, such as poor usability or unreliable content, but we understand that most of these points have been addressed in subsequent software upgrades.

Further work to develop sign-posting of potentially relevant content may expand the usage and therefore the benefits of CHIE. Negative perceptions or workload pressures might be outweighed if discovering unexpectedly useful data became a routine experience^29^.

What is the potential of CHIE to have an impact on major NHS cost pressures? We do not conclude that CHIE can significantly reduce hospital admissions or accelerate discharges. The efficiency savings in medicines reconciliation and test avoidance are largely already realised in HIOW, but this is a feasible target for other regions and countries implementing inter-organisational shared care records. Different configurations of shared records or alternative service designs may make reduction in admissions or expedited discharges feasible targets in other contexts. More extensive content from social care might support better service integration and lead to facilitated discharge decisions and processes.

### Limitations

Our small sample size meant that we were unable to draw meaningful conclusions about differentiation between specialties or professions or be certain that we had reached saturation.

The focus of this study was on hospital usage, not primary, community or social care. We were only able to interview physicians and nurses, not pharmacists.

## Conclusions

There is no question that the basic aim of information sharing between sectors to enable quick answers to clinical questions is ethically right and practically achievable. There are definitely efficiency benefits to be realised in medicines reconciliation, but the only substantial cost saving that seems replicable is the reduction in duplicate laboratory testing.

We encourage further work to explore how potentially relevant information for the case at hand can be automatically identified and pulled out from the mass of data so as to help clinicians quickly assemble the patient story and find unexpected gems of knowledge. The concept of “recommender systems” is mature and well-known in other fields^22^, but does not yet seem to had significant application within health informatics^22^.

## Authors contribution

The study was conceived by PS and PR. The interviews were performed by HN. The qualitative data analysis was conducted by EA. This paper was drafted by PS and EA and all authors contributed to revising it and approving the final text.

## Data Availability

The data consists of qualitative field notes and transcripts, which we do not plan to make available unless there is particular justification.

## Acknowledgements

We gratefully acknowledge the advice of our PPI representatives, the clinicians who gave their time to be interviewed and the other NHS staff who facilitated the interview recruitment.

## Appendix – Semi-structured interview outline

Thank you for your time today. May I first check that you are still willing to do this interview and for me to record it? The interview should take no more than 40 minutes, is that ok?

[If participant declines, thank and close the conversation. If they are willing to re-schedule, try to find a suitable time within the next week or so.]

As a reminder, the purpose of this study is to learn from your experiences of using the Hampshire Health Record, also known as the Care and Health Information Exchange (CHIE). In this survey we will use the name Hampshire Health Record (HHR).

[After each question 1-17: Pause, listen, probe. Where the script says HHR, use the full name “Hampshire Health Record”]

1. How often do you use the HHR in a typical day?
2. In what situations is the HHR most useful?
3. How often do you find what you need in the HHR? (approximate %)
4. What do you find most useful about the HHR?
5. What do you find most frustrating about the HHR?
6. How does the HHR help you when deciding whether to admit a patient?
7. How does the HHR help you in medicines reconciliation?
8. How does the HHR help you when deciding what tests to order for an admitted patient?
9. How does the HHR help you when discharging a patient?
10. Are there particular patient groups or disease types where the HHR is particularly useful?
11. Are there particular patient groups or disease types where the HHR is seldom useful?
12. Do you think that the HHR can reduce length of stay? How?
13. Do you think that the HHR has enabled quality improvements or cost savings? How?
14. Why do you think some clinicians make little use of the HHR?
15. Do you use the Summary Care Record, the TPP Viewer or the MIG?
16. In what situations is the Summary Care Record, the TPP Viewer or the MIG most useful?
17. Have you worked in organisations that do not have a shared record like the HHR? If so, what do you see as the main benefits of having a regional shared record?
18. On balance, do you think HHR is good for the citizens of Hampshire & IOW?

### Close

Would you like to make any other comments about HHR? [Pause, listen, probe.]

Would you be willing to participate in any follow-up research? [If so, confirm contact details.]

Thank you again for taking the time to do this interview. As a reminder, all interviews will be analysed anonymously and we will not identify you in any published report.

